# Regional variation in ADHD treatment and diagnosis in Denmark

**DOI:** 10.1101/2025.10.20.25338348

**Authors:** Heidi Sonne, Anton Pottegård, Anna Sofie Kjærgaard Hansen, Mickael Bech, Helene Kildegaard

## Abstract

**Introduction:** Attention-deficit/hyperactivity disorder (ADHD) treatment and diagnoses show marked international and regional variation, not fully explained by underlying morbidity. In Denmark, earlier studies reported substantial regional differences, raising concerns about inequities in access to care. We examined nationwide patterns of ADHD medication use and diagnosis in 2024 and assessed system-level factors potentially contributing to geographic variation.

**Methods:** We conducted a register-based cross-sectional study of all Danish residents aged 4-17 years in 2024, using data from the National Patient Register, National Prescription Registry, Civil Registration System, and administrative sources. Pharmacological treatment of ADHD was defined as at least one redeemed prescription for ADHD drugs, while ADHD diagnoses were defined as at least one hospital contact with International Classification of Diseases, 10th revision codes F90.x/F98.8. Prevalence estimates were calculated overall and stratified by subgroups of sex and age. Associations with municipal socioeconomic index, regional hospital waiting times, and private child psychiatrist capacity were evaluated using Spearman’s rank correlations and regression analyses.

**Results:** Among 4-17-year-olds, ADHD medication prevalence ranged sixfold across municipalities (9.6-58 per 1,000). ADHD diagnoses showed parallel patterns (15-74 per 1,000). Variation was largest among adolescents (more than sevenfold). Municipal socioeconomic status was weakly and inversely correlated with medication use (ρ = -0.20, p = 0.049), but not diagnoses. Regional waiting times and specialist capacity varied but showed no significant associations with the prevalence of either medication use or diagnoses.

**Conclusion:** Marked geographic variation in ADHD medication use and diagnoses persists in Denmark despite a uniform healthcare framework. Observed differences were only weakly related to socioeconomic context, specialist capacity, or waiting times, suggesting that the unwarranted variation is driven by other factors. Ensuring equitable access will require addressing both structural resources and local practice variation.

**Significant Outcomes:** ADHD medication and diagnosis prevalence among Danish children and adolescents in 2024 varied more than sixfold across municipalities.

Socioeconomic context, specialist capacity, and waiting times explained little of the observed geographic variation.

**Limitations:** Waiting time data were available only for hospital-based services, excluding the referral phase and private practices.

Specialist capacity was measured only for private psychiatrists under agreements with the public health insurance system, limiting coverage of capacity for public hospitals and private non-contracted services.

Regional-level analyses were constrained by the small number of regions (n=5), reducing statistical power and precision.

## Introduction

The prevalence of attention-deficit/hyperactivity disorder (ADHD) diagnoses has increased substantially over recent decades. International studies have documented marked regional variation in both the diagnosis and treatment of ADHD, including evidence from Norway, Sweden, and the United States.^1–3^ Such differences are only partly explained by underlying patient needs and may also reflect supply- and demand-side factors, including access to services and local medical practices.^4,5^

Geographic variation in healthcare utilization has been well documented for decades, often referred to as “small area variation.”^4^ These differences are typically regarded as unwarranted when they cannot be attributed to evidence-based guidelines or population health needs.^6^ Similar concerns apply to ADHD, where local practices and system-level factors may drive differential access to diagnostic and treatment services.

In Denmark, approximately 9% of children receive an ADHD diagnosis before age 18.^7^ Earlier Danish research has found substantial regional variation in incidence, with lower rates in the Southern Region, particularly on Funen, and higher rates in the Zealand Region.^8^ These findings point to unwarranted variation in care delivery and highlight potential inequalities in access to diagnosis and treatment.

This study provides updated nationwide estimates of geographic variation in ADHD treatment in Denmark in 2024. We further examine system-level factors that may drive these differences to assess whether regional variation reflects true population needs or unwarranted variation in care. By doing so, we aim to generate evidence that can inform strategies to promote more equitable access to ADHD treatment across the country.

## Methods

We conducted a nationwide register-based study to estimate the prevalence of diagnosed ADHD and ADHD medication use among Danish children and adolescents, and to examine selected regional and municipal indicators potentially associated with variation. Data were obtained from the Danish National Patient Register,^9^ the Danish National Prescription Registry,^10^ the Danish Civil Registration System,^11^ and public administrative data from the Ministry of the Interior and Health,^12^ as well as the Danish Health Data Authority.^13,14^

### Setting

Denmark has a universal, tax-funded healthcare system in which all residents are entitled to publicly financed healthcare. Access to diagnostic assessments, consultations, and hospital-based services is provided free of charge, while a fixed percentage copayment applies to outpatient prescription drugs.^15^ The system is organized into five regions responsible for managing hospital services, general practitioners, and private specialist care:^16^ The Capital Region of Denmark (∼1.9 million inhabitants), Region Zealand (∼0.8 million), the Region of Southern Denmark (∼1.2 million), the Central Denmark Region (∼1.4 million), and the North Denmark Region (∼0.6 million).^15,17^ These regions are subdivided into 98 municipalities, most of which have at least 20,000 inhabitants.^17^

Child and adolescent psychiatry in Denmark is organized as a specialized regional service.^16^ Access requires referral from a general practitioner, the municipality, or another authorized healthcare professional, after which referrals are assessed by the regional child and adolescent psychiatry services.^18,19^ Once a referral is accepted, the patient undergoes a standardized diagnostic assessment, followed by a treatment plan that may include pharmacological treatment. Dependent on symptom severity, complexity, and waiting time, patients may be referred to private practicing child and adolescent psychiatry clinics with a provider number,^20^ meaning that they are under agreements with the public health insurance system. In 2023, approximately 5,000 patients were treated by private child and adolescent psychiatrists with a provider number, compared with about 48,700 patients in hospital-based child and adolescent psychiatry.^20^ An unknown number of private child and adolescent psychiatrists without a provider number work outside the public reimbursement system, requiring full out-of-pocket payment by families.^20^

Diagnostic information from private practices without a provider number is not systematically reported to the Danish National Patient Register, while all prescriptions redeemed at pharmacies are mandatorily recorded in the Danish National Prescription Registry,^10^ enabling more complete monitoring of medication use compared with diagnostic data.

### Study population

The study population was all individuals aged 4–17 years residing in Denmark between January 1 and December 31, 2024. The population was obtained from the Danish Civil Registration System along with key demographic information, including date of birth, sex, and residential region and municipality. The age range was chosen from 4 years, as ADHD symptoms can be difficult to distinguish from normative behaviour before this age,^21^ and diagnosis before age 4 is rare.^22^

### Analyses

Prevalence estimates were calculated overall and stratified by sex and age groups (4-6, 7-12, and 13-17 years), region, and municipality. Age groups were chosen to align with key stages in the Danish educational system: kindergarten and pre-school (4-6 years), primary and middle school (7-12 years), and secondary school to early upper secondary education (13-17 years). For analyses stratified by municipality, residence as of December 31, 2024, was used to assign individuals.

Users of ADHD medication were defined as individuals who redeemed at least one prescription for methylphenidate (Anatomic Therapeutic Chemical (ATC) code: N06BA04 (62% of all defined daily doses filled in 2024 for age 0-17)^23^, lisdexamphetamine (N06BA12; 29%), atomoxetine (N06BA09; 6.7%), dexamphetamine (N06BA02; 1.4%), or guanfacine (C02AC02; 1.1%) between January 1 and December 31, 2024, identified using the Danish National Prescription Registry.^10^

The prevalence of ADHD diagnoses was defined as the proportion of individuals in the study population who had received a diagnosis before December 31, 2024, using all available lookback. An ADHD diagnosis was defined as at least one hospital contact, either inpatient or outpatient, recorded in the Danish National Patient Register with a diagnosis according to the International Classification of Diseases, 10th revision (ICD-10), codes F90.x or F98.8.^9,24^

To explore potential drivers of geographic variation, we included three types of regional and municipality-level indicators, capturing both demand-(1) and supply-side factors (2+3): 1) The municipal socioeconomic index, obtained from the Ministry of the Interior and Health, which estimates each municipality’s relative expenditure needs based on a range of indicators, including the proportion of adults not in employment and the number of residents with psychiatric conditions.^12^ Values above 1 indicate higher-than-average expenditure needs, while values below 1 indicate lower-than-average needs.^12^ 2) Waiting time in the hospital sector, obtained from the Danish Health Data Authority, defined as the period from the completion of diagnostic workup at the hospital to treatment initiation, or, if workup occurred outside the hospital system, from the receipt of referral to treatment start.^13^ 3) The number of private practicing child and adolescent psychiatrists with a provider number, obtained from the Danish Health Data Authority and expressed as full-time equivalent capacities (FTEs) per 100,000 inhabitants, where a full-time practice counted as one FTE and part-time practices typically counted as one-third FTE.^14^

Associations between municipal and regional indicators and the prevalence of ADHD diagnoses and medication use were assessed using Spearman’s rank correlation coefficients (ρ). For the municipal socioeconomic index, we additionally fitted population-weighted linear regression models with prevalence proportion as the outcome. For the regional indicators (waiting time and psychiatrist capacity), associations were further examined using simple linear regressions based on regional prevalence estimates.

All analyses were performed using R Studio version 4.4.3.

## Results

In 2024, the prevalence of ADHD medication use among Danish children aged 4-17 years varied substantially across municipalities, ranging from 9.6 to 17 per 1,000 in municipalities on Funen to 30 to 58 per 1,000 in municipalities in the North Denmark Region and the Central Denmark Region (**Figure 1A**) (**Supplementary Table 1**). ADHD diagnoses followed a similar pattern, with the lowest prevalence (15 to 26 per 1,000) in municipalities on Funen and the highest (32 to 74 per 1,000) in municipalities in the North Denmark Region and the Central Denmark Region (**Figure 1B**).

**Figure 1.**
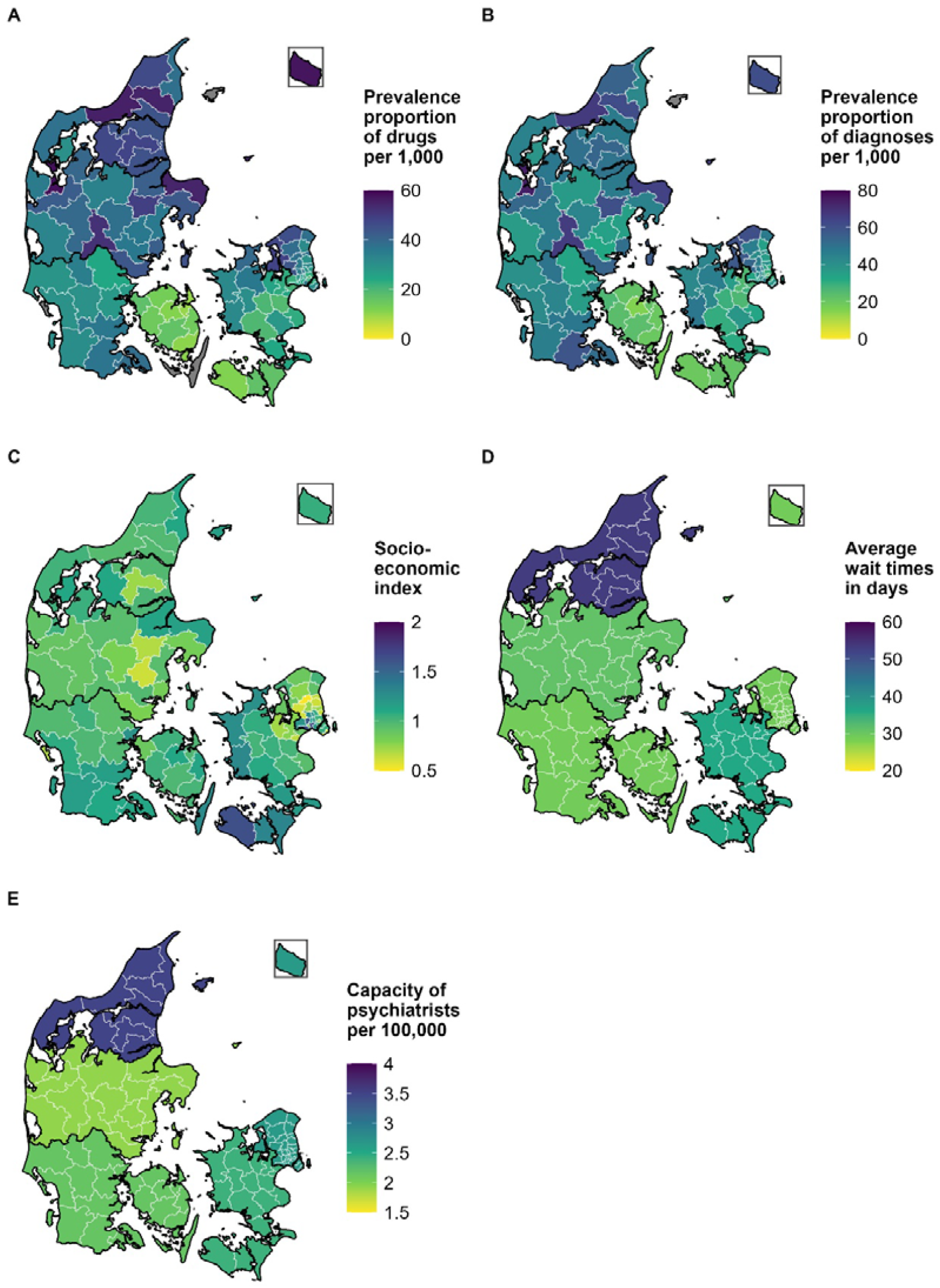
Prevalence of ADHD medication use and ADHD diagnoses among children and adolescents aged 4-17 years in Denmark, 2024. (A) Municipal prevalence of ADHD medication use in 2024 defined as ≥1 redeemed prescription for methylphenidate (N06BA04), atomoxetine (N06BA09), dexamphetamine (N06BA02), lisdexamphetamine (N06BA12), or guanfacine (C02AC02), per 1,000 inhabitants aged 4-17 years. (B) Municipal prevalence of ADHD diagnoses in 2024 defined as ≥1 inpatient or outpatient hospital contact in the Danish National Patient Register with ICD-10 F90.x or F98.8, per 1,000 inhabitants aged 4-17 years. (C) Municipal socioeconomic index obtained from the Ministry of the Interior and Health (values >1 indicate higher-than-average needs). (D) Regional waiting times as the mean days from completed diagnostic workup or referral to treatment initiation in the 3^rd^ quarter of 2024, obtained from the Danish Health Data Authority. (E) Regional capacity of private practicing psychiatrists with a provider number, expressed as full-time equivalents per 100,000 inhabitants aged 4-17 years.

When stratified by age, variation in medication use was lowest among children aged 4-6 years (1.2 to 4.7 per 1,000 across municipalities with reportable numbers), higher among children aged 7-12 years (8.9 to 62 per 1,000), and highest among adolescents aged 13-17 years (12 to 94 per 1,000) (**Supplementary Figure 1**). ADHD diagnoses showed the same age-dependent pattern, with the widest geographic variation among adolescents (22 to 113 per 1,000), somewhat narrower variation in children aged 7-12 years (15 to 74 per 1,000), and low rates in children aged 4-6 years (3.1 to 10 per 1,000) (**Supplementary Figure 2**).

When stratified by sex, ADHD medication use was consistently higher among boys than girls. For girls, municipal prevalence ranged from 7.6 to 46 per 1,000, while for boys the range was 10 to 73 per 1,000 (**Supplementary Figure 3**). Diagnostic prevalence showed a similar sex difference, ranging from 9.3 to 48 per 1,000 among girls and 21 to 100 per 1,000 among boys (**Supplementary Figure 4**).

Socioeconomic index values varied across municipalities with no clear regional pattern (**Figure 1C**) (**Supplementary Table 2**). ADHD medication use was slightly higher in municipalities with lower relative expenditure needs (ρ = -0.20, p = 0.049), whereas no significant association was found for ADHD diagnoses (ρ = -0.04, p = 0.68) (**Supplementary Table 3**). The association between socioeconomic index and medication was stronger for girls (ρ = -0.29, p = 0.005) than for boys (ρ = -0.15, p = 0.15). Results from regression analyses were similar to the correlation findings, with a statistically significant association for medication use (β = -0.013, p = 0.027) and no association for ADHD diagnoses (β = -0.004, p = 0.51) (**Supplementary Table 4**).

Regions with higher medication use also tended to report longer waiting times, most notably in the North Denmark Region (prevalence 45 per 1,000; average waiting time 53 days) compared to the Capital Region and Region of Southern Denmark, where medication prevalence was lower (30 and 23 per 1,000) and waiting times shorter (29 days) (**Figure 1D**). However, waiting times were not statistically significantly correlated with either medication use (ρ = 0.62, p = 0.27) or diagnoses (ρ = 0.36, p = 0.55).

Capacity in private child and adolescent psychiatry with provider numbers also varied geographically, ranging from 1.9 per 100,000 in the Central Denmark Region to 3.5 per 100,000 in the North Denmark Region, but was likewise not significantly associated with medication use (ρ = 0.40, p = 0.50) or ADHD diagnoses (ρ = 0.30, p = 0.62) (**Figure 1E**).

## Discussion

Our nationwide register-based study found marked geographic variation in ADHD diagnosis and medication use among Danish children in 2024, with prevalences differing more than sixfold across municipalities. Municipal socioeconomic status, private specialist capacity, and public hospital waiting times explained very little of these differences. Importantly, such variation should not be interpreted as low prevalence being inherently preferable or high prevalence being undesirable. Rather, the concern is that the observed magnitude of variation appears too great given the presence of uniform clinical guidelines across the country and the lack of clear explanatory factors in our data.

Similar variations in ADHD diagnoses have been reported previously in Denmark, with high diagnostic rates in Region Zealand and low rates in the Region of Southern Denmark, especially on Funen.^8^ Comparable findings have been observed in Norway, where geographic differences in diagnostic rates greatly exceeded variation in underlying ADHD symptom levels, indicating that morbidity alone cannot account for geographic patterns.^2^ Likewise, regional variability in ADHD within a universal health system has been documented in Spain,^25^ suggesting that structural uniformity does not eliminate local-level differences.

We observed no consistent socioeconomic gradient in ADHD diagnoses but found a weak negative correlation between the socioeconomic index and medication use, suggesting slightly higher medication use in municipalities with lower relative expenditure needs. Although previous studies have reported higher ADHD prevalence in disadvantaged areas,^26,27^ and more consistent pharmacotherapy among children from wealthier families,^28^ we observed no clear socioeconomic gradient. Earlier Danish studies also point in different directions: one study found considerable geographic clustering of ADHD incidence and medication use, but no significant associations with socioeconomic or municipal-level factors,^29^ while another showed that regional prescribing differences were particularly pronounced among children facing social adversity, reflecting strong social gradients in some regions but not in others.^30^ Norwegian data show that socioeconomic indicators are more strongly associated with service contact rates than with ADHD prevalence once children access mental health services.^1^ They suggest that inequalities act mainly at the access stage, not at the diagnostic stage. Conceptual frameworks of healthcare access further highlight that such differences may occur at multiple stages of the care pathway, from the perception of needs to health care seeking, reaching, and engagement, and not only at the point of diagnosis.^31^

Specialist availability may also contribute to variation. The North Denmark Region had both the highest private specialist capacity and the highest prevalence, but also the longest public-sector waiting times. Results from the United States show that higher numbers of relevant physicians are associated with greater ADHD diagnosis and medication rates.^32^ This suggests that supply-sensitive care may be a partial driver: higher availability of specialist services may increase detection and diagnosis, even when demand is constant.^6^ However, the lack of statistical associations in our study, together with high prevalence in parts of Zealand despite relatively low capacity, points to additional drivers such as parental demand or local diagnostic culture. Research from Norway has shown inter-clinician variation in diagnostic thresholds,^33^ and a similar factor could plausibly influence Danish practice despite uniform national guidelines. Evidence from Denmark further indicates that specialist physicians differ considerably in their prescribing behaviour across hospitals, and that such variation directly affects the probability that children receive pharmacological treatment.^34^ Such preference-sensitive care, driven by differing professional judgments, remains a likely contributor.^6^

Educational system incentives may also play a role. In some municipalities, access to special educational support requires a formal diagnosis, which can create incentives for parents, schools, and professionals to seek diagnostic assessments. A recent qualitative study highlights how ADHD diagnoses are used institutionally in some Danish primary schools as a prerequisite for categorizing children with emotional and behavioural difficulties and for allocating support services.^35^ Such mechanisms could plausibly contribute to municipal differences if the reliance on diagnostic categorization varies across local school systems.

Community-level dynamics may further shape ADHD prevalence. In autism, proximity to diagnosed children increases the likelihood of receiving a diagnosis, likely through heightened awareness and parent-to-parent information exchange.^36^ Similar processes may influence ADHD patterns.

### Future research

Beyond the factors examined in this study, other demand- and supply-side mechanisms may also influence regional variation in ADHD treatment. On the demand side, parental health-seeking behaviour, school-level incentives, and local awareness networks may shape both referral patterns and treatment uptake. On the supply side, differences in distance to care, the distribution of private practicing and public hospital-based child and adolescent psychiatrists, regional prioritization of resources, and varying professional norms, including potential differences between private and hospital-based diagnostic practices, could contribute. Future research should therefore combine register data with qualitative and survey-based approaches to capture these dimensions and assess how such mechanisms interact with structural factors to generate the observed variation.

### Strengths and Limitations

Key strengths include the use of nationwide registry data, ensuring complete population coverage, and eliminating selection bias. Prescription data include all community pharmacy dispensations, regardless of prescriber or sector, and diagnostic data cover both public and private providers with a provider number.

Several limitations should be noted. First, waiting time from diagnostic assessment until treatment initiation was available only for hospital-based child and adolescent psychiatry, and we did not have data on waiting time at public hospitals from referral to diagnostic assessment. Current expected waiting times reported by private practicing psychiatrists with provider numbers can be obtained,^37^ but historical data are lacking.

Second, specialist capacity data were available only for private practicing psychiatrists with a provider number. Information on public hospital-based capacity is available in the Health Workforce Mobility Register,^38^ which was not obtained for this study, and publicly accessible data on hospital capacity divided by region extends only until 2022.^39^ Moreover, diagnostic information from private practices without provider numbers is not systematically reported to the Danish National Patient Register, limiting the completeness of the number of ADHD diagnoses.

Third, prevalence was defined as at least one prescription in 2024, which may overestimate active treatment rates, though relative geographic patterns are unlikely to change. Third, statistical associations at the regional level were strongly limited by the small number of regions (n = 5), which reduced power and increased uncertainty around effect estimates. Additionally, the organisation of child and adolescent mental health services varies geographically, with some areas served by multiple units and others by only one. In the latter case, a single department can account for a large proportion of diagnoses in the region, potentially amplifying local practice effects.

Finally, we lacked data on symptom severity, parental demand, local clinical practice norms, and individual-level socioeconomic background, all of which may be important explanatory factors.

## Conclusion

Large geographic differences in ADHD diagnosis and medication use persist in Denmark despite a uniform healthcare framework. These differences appear only weakly related to specialist capacity, waiting times, and socioeconomic context, suggesting that variation might be driven by other factors. Reducing inequities will likely require a multifaceted approach that addresses both resource distribution and local practice variation, ensuring equitable and consistent pathways to diagnosis and treatment.

## Data Availability

Individual-level data cannot be shared by the authors owing to Danish data protection regulations. De-identified data can be made available for authorized researchers after application with a relevant aim to Forskerservice at the Danish Health Data Authority.

