## Supplementary Material for "Regional variation in ADHD treatment and diagnosis in Denmark"

**Supplementary Table 1.** Prevalence of ADHD medication use and ADHD diagnoses among children and adolescents aged 4-17 years in Denmark, 2024. Estimates are shown per 1,000 inhabitants at national, regional, and municipal levels.

| Geographic entity | Prevalence proportion of drugs per 1,000 | Prevalence proportion of diagnosis per 1,000 |
| --- | --- | --- |
| National | 32 | 39 |
| The Capital Region of Denmark | 30 | 38 |
| Albertslund | 29 | 40 |
| Allerød | 38 | 41 |
| Ballerup | 35 | 48 |
| Bornholm | 57 | 61 |
| Brøndby | 25 | 40 |
| Dragør | 36 | 40 |
| Egedal | 44 | 53 |
| Fredensborg | 31 | 34 |
| Frederiksberg | 19 | 23 |
| Frederikssund | 48 | 58 |
| Furesø | 35 | 46 |
| Gentofte | 28 | 31 |
| Gladsaxe | 29 | 35 |
| Glostrup | 28 | 46 |
| Gribskov | 45 | 61 |
| Halsnæs | 47 | 60 |
| Helsingør | 38 | 47 |
| Herlev | 37 | 48 |
| Hillerød | 38 | 46 |
| Hvidovre | 35 | 43 |
| Høje-Taastrup | 22 | 33 |
| Hørsholm | 38 | 42 |
| Ishøj | 24 | 37 |
| København | 23 | 30 |
| Lyngby-Taarbæk | 33 | 39 |
| Rudersdal | 30 | 36 |
| Rødovre | 27 | 34 |
| Tårnby | 30 | 37 |
| Vallensbæk | 24 | 33 |
| Region Zealand | 25 | 33 |
| Faxe | 28 | 38 |
| Greve | 21 | 28 |
| Guldborgsund | 17 | 21 |
| Holbæk | 36 | 46 |
| Kalundborg | 32 | 43 |
| Køge | 19 | 26 |
| Lejre | 25 | 35 |
| Lolland | 12 | 20 |
| Næstved | 19 | 30 |
| Odsherred | 39 | 44 |
| Ringsted | 20 | 29 |
| Roskilde | 23 | 26 |
| Slagelse | 32 | 49 |
| Solrød | 24 | 29 |
| Sorø | 30 | 43 |
| Stevns | 23 | 33 |
| Vordingborg | 24 | 31 |
| The Region of Southern Denmark | 23 | 34 |
| Assens | 14 | 24 |
| Billund | 34 | 49 |
| Esbjerg | 30 | 42 |
| Fanø | N < 10 | N < 10 |
| Fredericia | 27 | 42 |
| Faaborg-Midtfyn | 16 | 24 |
| Haderslev | 36 | 48 |
| Kerteminde | 10 | 22 |
| Kolding | 28 | 36 |
| Langeland | N < 10 | 18 |
| Middelfart | 17 | 26 |
| Nordfyns | 13 | 22 |
| Nyborg | 14 | 20 |
| Odense | 11 | 15 |
| Svendborg | 11 | 20 |
| Sønderborg | 38 | 51 |
| Tønder | 31 | 46 |
| Varde | 29 | 40 |
| Vejen | 30 | 44 |
| Vejle | 24 | 34 |
| Ærø | N < 10 | N < 10 |
| Aabenraa | 37 | 59 |
| The Central Denmark Region | 39 | 43 |
| Favrskov | 49 | 60 |
| Hedensted | 44 | 54 |
| Herning | 39 | 47 |
| Holstebro | 42 | 53 |
| Horsens | 36 | 32 |
| Ikast-Brande | 52 | 66 |
| Lemvig | 35 | 45 |
| Norddjurs | 55 | 65 |
| Odder | 42 | 51 |
| Randers | 42 | 47 |
| Ringkøbing-Skjern | 33 | 36 |
| Samsø | 45 | 57 |
| Silkeborg | 38 | 44 |
| Skanderborg | 36 | 35 |
| Skive | 40 | 48 |
| Struer | 58 | 74 |
| Syddjurs | 43 | 45 |
| Viborg | 33 | 37 |
| Aarhus | 36 | 35 |
| The North Denmark Region | 45 | 50 |
| Brønderslev | 55 | 61 |
| Frederikshavn | 38 | 43 |
| Hjørring | 46 | 55 |
| Jammerbugt | 55 | 67 |
| Læsø | N < 10 | N < 10 |
| Mariagerfjord | 45 | 50 |
| Morsø | 30 | 38 |
| Rebild | 48 | 54 |
| Thisted | 38 | 45 |
| Vesthimmerlands | 47 | 50 |
| Aalborg | 44 | 47 |

**Supplementary Table 2.** Regional and municipal indicators in Denmark, 2024. Indicators include municipal socioeconomic index (values >1 indicate higher-than-average needs), average regional waiting times in public child and adolescent psychiatry (days), and regional capacity of private practicing child and adolescent psychiatrists with a provider number (full-time equivalents per 100,000 inhabitants aged 4-17 years).

| Geographic entity | Socioeconomic index | Average wait time (days) | Capacity of private psychiatrists (FTE) per 100,000 |
| --- | --- | --- | --- |
| The Capital Region of Denmark |  | 29 | 2.6 |
| Albertslund | 1.56 |  |  |
| Allerød | 0.51 |  |  |
| Ballerup | 1.27 |  |  |
| Bornholm | 1.06 |  |  |
| Brøndby | 1.66 |  |  |
| Dragør | 0.62 |  |  |
| Egedal | 0.61 |  |  |
| Fredensborg | 0.92 |  |  |
| Frederiksberg | 0.76 |  |  |
| Frederikssund | 0.89 |  |  |
| Furesø | 0.74 |  |  |
| Gentofte | 0.64 |  |  |
| Gladsaxe | 1.08 |  |  |
| Glostrup | 1.18 |  |  |
| Gribskov | 0.87 |  |  |
| Halsnæs | 1.02 |  |  |
| Helsingør | 1.02 |  |  |
| Herlev | 1.25 |  |  |
| Hillerød | 0.77 |  |  |
| Hvidovre | 1.19 |  |  |
| Høje-Taastrup | 1.22 |  |  |
| Hørsholm | 0.64 |  |  |
| Ishøj | 1.62 |  |  |
| København | 1.04 |  |  |
| Lyngby-Taarbæk | 0.73 |  |  |
| Rudersdal | 0.62 |  |  |
| Rødovre | 1.22 |  |  |
| Tårnby | 1.06 |  |  |
| Vallensbæk | 0.82 |  |  |
| Region Zealand |  | 36 | 2.4 |
| Faxe | 1.02 |  |  |
| Greve | 0.90 |  |  |
| Guldborgsund | 1.32 |  |  |
| Holbæk | 1.07 |  |  |
| Kalundborg | 1.26 |  |  |
| Køge | 1.02 |  |  |
| Lejre | 0.72 |  |  |
| Lolland | 1.63 |  |  |
| Næstved | 1.09 |  |  |
| Odsherred | 1.25 |  |  |
| Ringsted | 1.05 |  |  |
| Roskilde | 0.78 |  |  |
| Slagelse | 1.30 |  |  |
| Solrød | 0.65 |  |  |
| Sorø | 1.02 |  |  |
| Stevns | 0.95 |  |  |
| Vordingborg | 1.19 |  |  |
| The Region of Southern Denmark |  | 29 | 2.2 |
| Assens | 1.04 |  |  |
| Billund | 1.04 |  |  |
| Esbjerg | 1.08 |  |  |
| Fanø | 0.69 |  |  |
| Fredericia | 1.23 |  |  |
| Faaborg-Midtfyn | 0.99 |  |  |
| Haderslev | 1.16 |  |  |
| Kerteminde | 0.99 |  |  |
| Kolding | 0.98 |  |  |
| Langeland | 1.28 |  |  |
| Middelfart | 0.87 |  |  |
| Nordfyns | 1.04 |  |  |
| Nyborg | 1.13 |  |  |
| Odense | 1.11 |  |  |
| Svendborg | 1.01 |  |  |
| Sønderborg | 1.12 |  |  |
| Tønder | 1.17 |  |  |
| Varde | 0.97 |  |  |
| Vejen | 0.99 |  |  |
| Vejle | 0.94 |  |  |
| Ærø | 0.99 |  |  |
| Aabenraa | 1.10 |  |  |
| The Central Denmark Region |  | 31 | 1.9 |
| Favrskov | 0.68 |  |  |
| Hedensted | 0.80 |  |  |
| Herning | 0.90 |  |  |
| Holstebro | 0.89 |  |  |
| Horsens | 0.97 |  |  |
| Ikast-Brande | 0.95 |  |  |
| Lemvig | 0.91 |  |  |
| Norddjurs | 1.15 |  |  |
| Odder | 0.83 |  |  |
| Randers | 1.13 |  |  |
| Ringkøbing-Skjern | 0.92 |  |  |
| Samsø | 1.03 |  |  |
| Silkeborg | 0.81 |  |  |
| Skanderborg | 0.62 |  |  |
| Skive | 1.05 |  |  |
| Struer | 1.04 |  |  |
| Syddjurs | 0.83 |  |  |
| Viborg | 0.95 |  |  |
| Aarhus | 0.95 |  |  |
| The North Denmark Region |  | 53 | 3.5 |
| Brønderslev | 0.98 |  |  |
| Frederikshavn | 1.11 |  |  |
| Hjørring | 1.01 |  |  |
| Jammerbugt | 0.96 |  |  |
| Læsø | 1.16 |  |  |
| Mariagerfjord | 1.00 |  |  |
| Morsø | 1.11 |  |  |
| Rebild | 0.73 |  |  |
| Thisted | 1.03 |  |  |
| Vesthimmerlands | 1.11 |  |  |
| Aalborg | 0.97 |  |  |

**Supplementary Table 3.** Spearman’s rank correlation coefficients between prevalence of ADHD medication use or ADHD diagnoses and regional/municipal indicators in Denmark, 2024. Results are shown for children and adolescents aged 4-17 years overall and stratified by sex.

| **Sex** | **Variable X** | **Variable Y** | **N** | **Rho** | **P-value** |
| --- | --- | --- | --- | --- | --- |
| Both | Socioeconomic index | Drugs | 94 | -0,20 | 0,049 |
|  |  | Diagnosis | 95 | -0,042 | 0,68 |
|  | Capacity per 100,000 | Drugs | 5 | 0,40 | 0,50 |
|  |  | Diagnosis | 5 | 0,30 | 0,62 |
|  | Wait times | Drugs | 5 | 0,62 | 0,27 |
|  |  | Diagnosis | 5 | 0,36 | 0,55 |
| Girls | Socioeconomic index | Drugs | 93 | -0,29 | 0,005 |
|  |  | Diagnosis | 93 | -0,055 | 0,60 |
|  | Capacity per 100,000 | Drugs | 5 | 0,40 | 0,50 |
|  |  | Diagnosis | 5 | 0,40 | 0,50 |
|  | Wait times | Drugs | 5 | 0,62 | 0,27 |
|  |  | Diagnosis | 5 | 0,62 | 0,27 |
| Boys | Socioeconomic index | Drugs | 94 | -0,15 | 0,15 |
|  |  | Diagnosis | 95 | -0,027 | 0,80 |
|  | Capacity per 100,000 | Drugs | 5 | 0,30 | 0,62 |
|  |  | Diagnosis | 5 | 0,30 | 0,62 |
|  | Wait times | Drugs | 5 | 0,36 | 0,55 |
|  |  | Diagnosis | 5 | 0,36 | 0,55 |

**Supplementary Table 4.** Results from linear regression analyses of the association between prevalence of ADHD medication use or ADHD diagnoses and regional/municipal indicators in Denmark, 2024. Analyses are presented for children and adolescents aged 4-17 years overall and stratified by sex. Municipal-level analyses are population-weighted, whereas regional-level analyses are unweighted.

| **Sex** | **Variable X** | **Variable Y** | **Estimate (95% CI)** | **P-value** |
| --- | --- | --- | --- | --- |
| Both | Socioeconomic index | Drugs | -0,013 (-0,024; -0,0015) | 0,027 |
|  |  | Diagnosis | -0,0041 (-0,017; 0,0084) | 0,52 |
|  | Capacity per 100,000 | Drugs | 0,0082 (-0,016; 0,032) | 0,36 |
|  |  | Diagnosis | 0,0072 (-0,010; 0,024) | 0,27 |
|  | Wait times | Drugs | 0,00061 (-0,00067; 0,0019) | 0,23 |
|  |  | Diagnosis | 0,00050 (0,00039; 0,0014) | 0,17 |
| Girls | Socioeconomic index | Drugs | -0,012 (-0,020; 0,0036) | 0,0055 |
|  |  | Diagnosis | -0,0049 (-0,014; 0,0040) | 0,28 |
|  | Capacity per 100,000 | Drugs | 0,0029 (-0,0056; 0,011) | 0,36 |
|  |  | Diagnosis | 0,0028 (-0,0028; 0,0084) | 0,21 |
|  | Wait times | Drugs | 0,00042 (-0,00053; 0,0014) | 0,26 |
|  |  | Diagnosis | 0,00036 (-0,00031; 0,0010) | 0,19 |
| Boys | Socioeconomic index | Drugs | -0,013 (-0,028; 0,0013) | 0,073 |
|  |  | Diagnosis | -0,0030 (-0,020; 0,013) | 0,72 |
|  | Capacity per 100,000 | Drugs | 0,0053 (-0,010; 0,021) | 0,36 |
|  |  | Diagnosis | 0,0044 (-0,0071; 0,016) | 0,31 |
|  | Wait times | Drugs | 0,00078 (-0,00082; 0,0024) | 0,22 |
|  |  | Diagnosis | 0,00064 (-0,00050; 0,0018) | 0,17 |

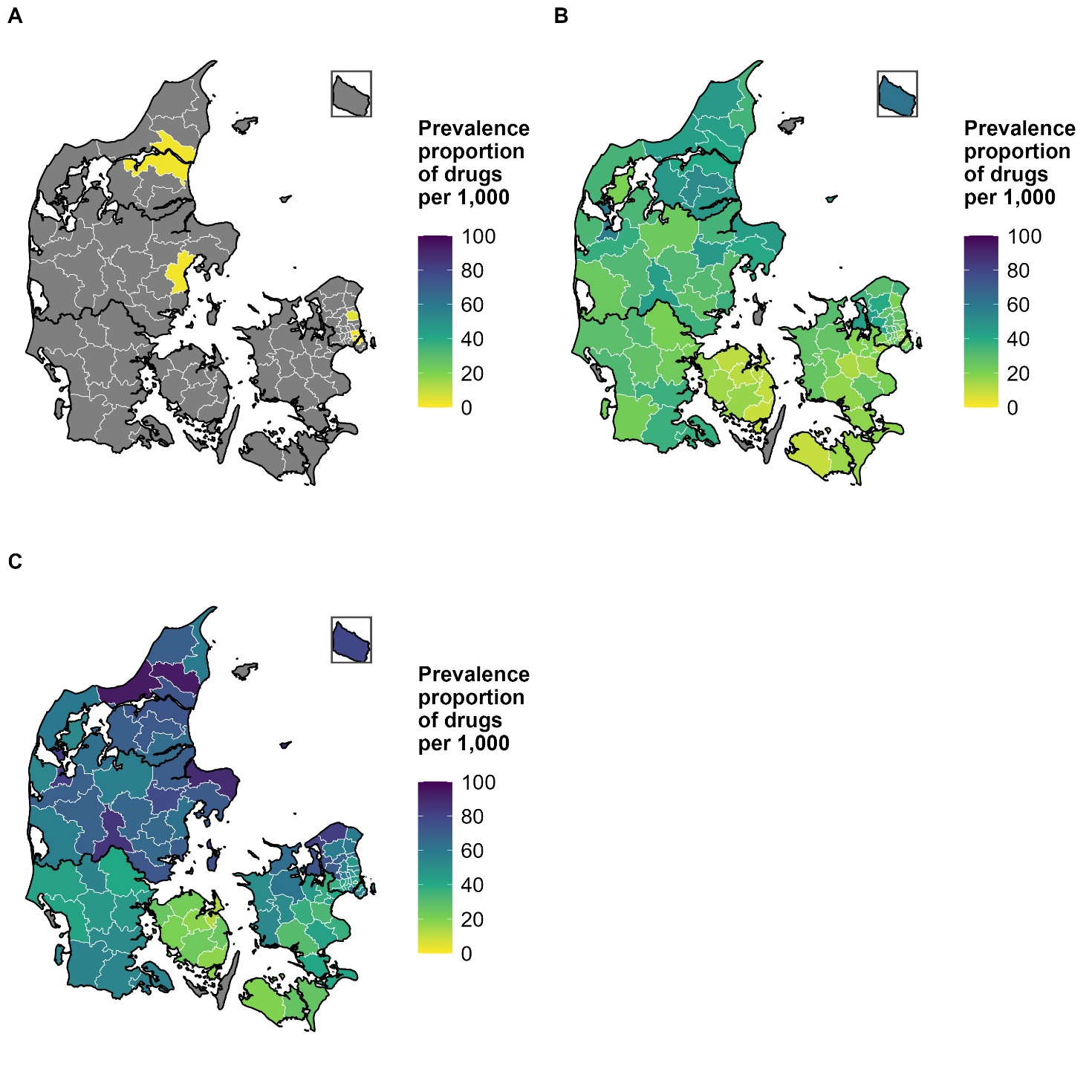

**Supplementary Figure 1.** Municipal prevalence of ADHD medication use per 1,000 inhabitants among children and adolescents aged 4-17 years in Denmark, 2024, stratified by age group.

(A) Children aged 4-6 years.
(B) Children aged 7-12 years.
(C) Adolescents aged 13-17 years.

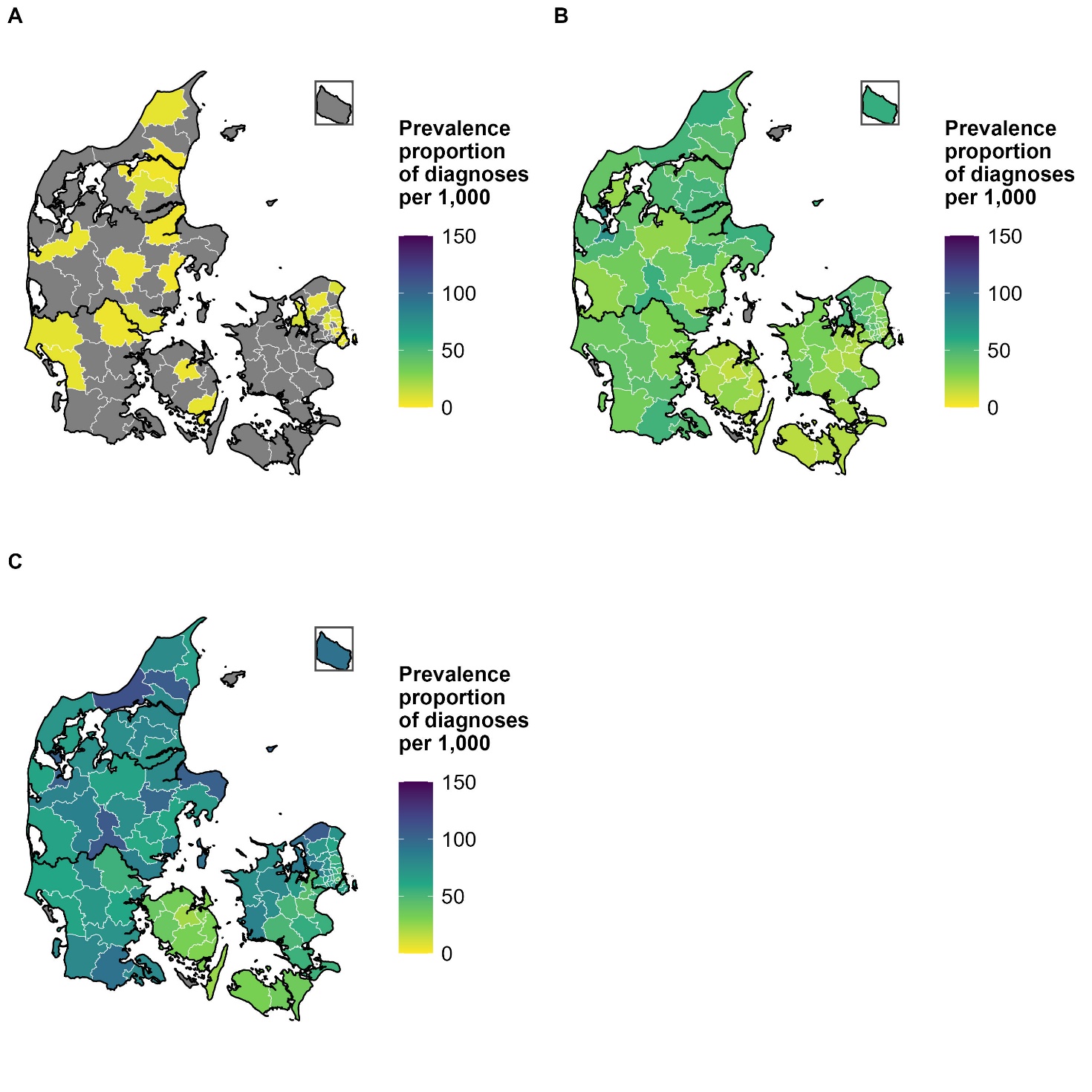

**Supplementary Figure 2.** Municipal prevalence of ADHD diagnoses per 1,000 inhabitants among children and adolescents aged 4-17 years in Denmark, 2024, stratified by age group.

(A) Children aged 4-6 years.
(B) Children aged 7-12 years.
(C) Adolescents aged 13-17 years.

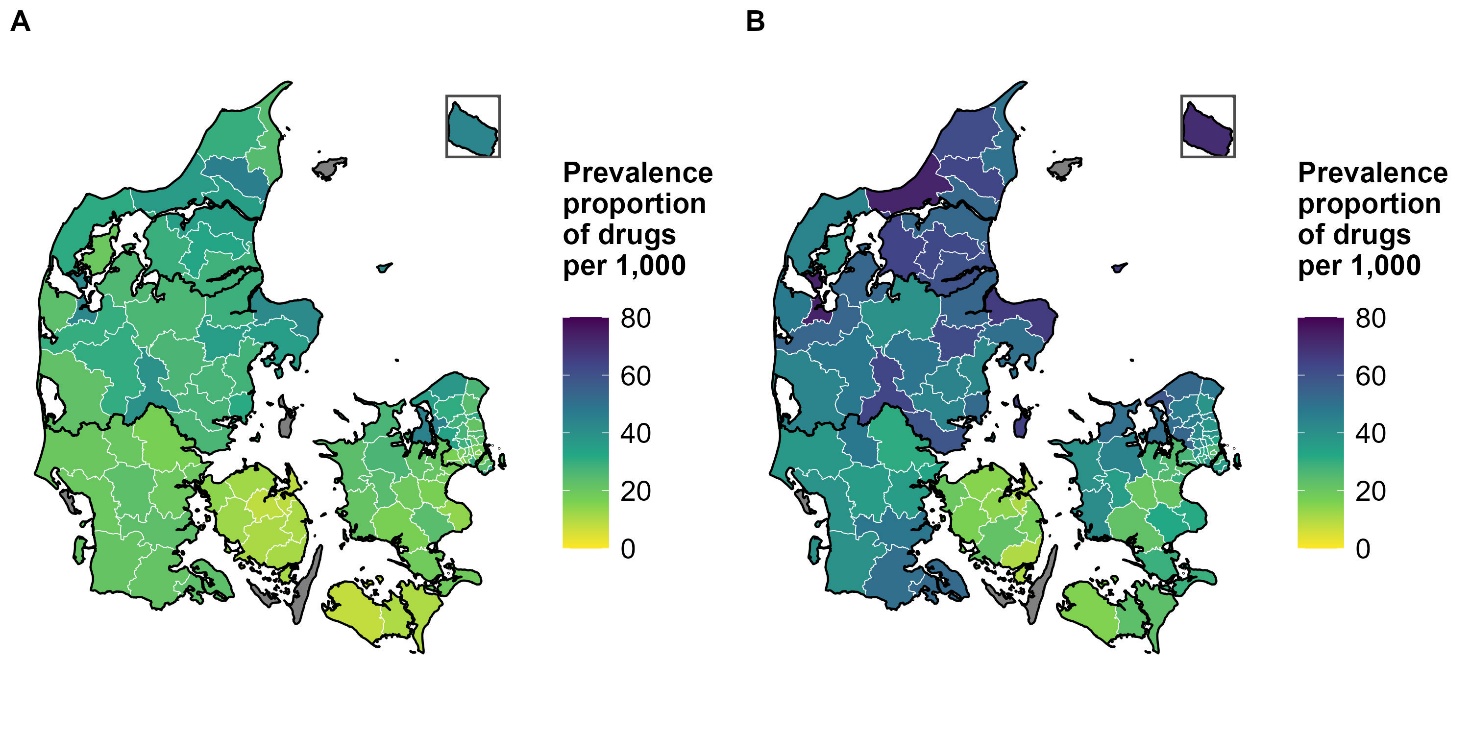

**Supplementary Figure 3.** Municipal prevalence of ADHD medication use per 1,000 inhabitants among children and adolescents aged 4-17 years in Denmark, 2024, stratified by sex.

(A) Girls.
(B) Boys.

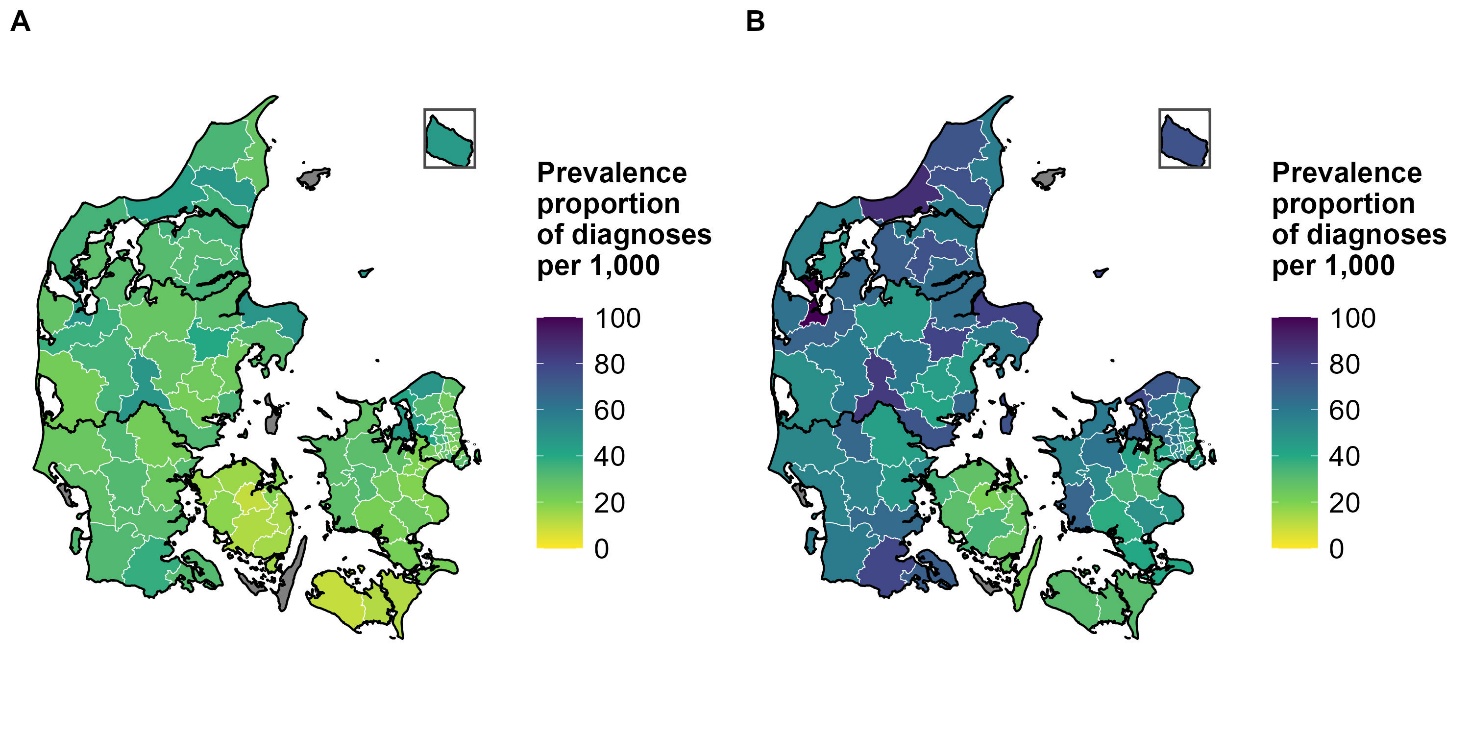

**Supplementary Figure 4.** Municipal prevalence of ADHD diagnoses per 1,000 inhabitants among children and adolescents aged 4-17 years in Denmark, 2024, stratified by sex.

(A) Girls.
(B) Boys.
